# Validated Synthetic Data Generation from a Multicenter Spine Surgery Registry: Methodology and Benchmark

**DOI:** 10.64898/2026.04.07.26350316

**Authors:** Vincent Challier, Clément Jacquemin, Bassel Diebo, Nassim Dehouche, Anton Denisov, Joseph Cristini, Mathieu Campana, Jean-Etienne Castelain, Guillaume Lonjon, Virginie Lafage, Soufiane Ghailane, SpineDAO Collaborative Group

**Affiliations:** Hôpital Privé du dos Francheville, Périgueux, France; Department of Orthopaedic Surgery, Brown University, Providence, RI, USA; Department of Orthopaedic Surgery, Lenox Hill Hospital, New York, NY, USA; Business Administration Division, Mahidol University International College, Salaya, Thailand; Department of Orthopedic Surgery, University Hospital Mollet, Barcelona, Spain; Biostatistics Division, The Taylor Collaboration, San Francisco, CA; Department of Neurosurgery, Hôpital Clairval, Marseille, France; Department of Spine Surgery, Clinique Orthosud, Saint Jean de Védas, France

**Keywords:** synthetic data, spine surgery, sacroiliac joint fusion, GaussianCopula, privacy-preserving machine learning, blockchain, registry, SpineBase

## Abstract

**Background:** Synthetic data have emerged as a complementary strategy for secondary use of clinical registries, enabling data sharing without patient-level exposure. In spine surgery, multicenter data sharing is constrained by institutional governance and patient privacy regulations. Validated synthetic data generation may enable broader access to surgical outcomes data for artificial intelligence development without compromising patient confidentiality.

**Objective:** To describe and benchmark a three-domain validated synthetic data pipeline applied to a multicenter, tokenized spine surgery registry (SpineBase), and to establish a reproducible certification framework for synthetic spine surgery datasets.

**Methods:** We extracted 125 sacroiliac joint fusion cases from the SpineBase registry (SIBONE study, IRB-SOFCOT approval Ref. 14–2025; CNIL MR-004 Ref. 2234503 v 0). A GaussianCopula generative model was trained on 52 structured variables spanning demographics, preoperative assessments, operative details, and longitudinal outcomes at 3, 6, 12, and 24 months. Synthetic datasets of 100, 1,000, and 10,000 patients were generated. Validation followed a three-domain framework: (1) fidelity, assessed by Kolmogorov-Smirnov tests and Jensen-Shannon divergence; (2) utility, assessed by train-on-synthetic, test-on-real (TSTR) methodology; and (3) privacy, assessed by nearest-neighbor distance ratio (NNDR), membership inference attack, and k-anonymity proxy.

**Results:** All three validation gates passed. Fidelity: mean KS p-value 0.52 (threshold >0.05). Privacy: NNDR >1.0 in 98.9% of synthetic records; membership inference AUROC 0.57. Utility: 12-month Oswestry Disability Index prediction yielded Pearson r = 0.29, consistent with expected attenuation at N = 125. A SHA-256 cryptographic hash of each certified dataset was anchored on the Solana blockchain for immutable provenance.

**Conclusions:** A validated, blockchain-anchored synthetic data pipeline for spine surgery registries is technically feasible and meets current publication-standard criteria for fidelity and privacy. Utility metrics scale with registry size, creating a direct incentive for multicenter data contribution. This framework provides a reproducible methodology for synthetic data certification in spine surgery research, and establishes certified synthetic datasets as a privacy-native substrate for expert-annotation pipelines — as demonstrated in the companion Spine Reviews study.

## INTRODUCTION

Modern spine surgery generates substantial volumes of structured clinical data — preoperative assessments, operative details, implant parameters, and longitudinal patient-reported outcomes — that collectively represent one of the richest phenotyped datasets in musculoskeletal medicine. Yet this data remains predominantly siloed within individual institutions, accessible neither for multicenter outcomes research nor for the large-scale artificial intelligence development that has transformed adjacent fields.^[1,2]^

The barriers to data sharing in spine surgery are well characterized: institutional governance requirements, heterogeneity of electronic health record systems, patient privacy regulations (HIPAA in the United States, GDPR in Europe), and the absence of agreed data standards. Several cooperative registries have addressed the governance problem through federated architectures or data use agreements, but the computational infrastructure for large-scale collaborative AI development remains immature.^[3,4]^

Synthetic data have attracted increasing attention as a complementary strategy. By training generative models on real clinical data and releasing statistically equivalent synthetic records, investigators may enable AI development and outcomes research without direct exposure of patient-level information. Greenberg and colleagues demonstrated in 2023 that EHR-derived synthetic data for cervical and lumbar fusion could preserve descriptive characteristics and predictive performance across a 15-hospital cohort, providing the strongest published proof of concept for this approach in spine surgery.^[5]^

Beyond privacy-preserving secondary use, synthetic data offer a second utility: serving as the substrate for expert-annotated AI training datasets. Synthetic patient vignettes can be distributed globally to verified specialists without institutional governance restrictions, enabling large-scale annotation at a fraction of traditional cost. This approach was demonstrated in a companion study by the SpineDAO Collaborative Group.^[24]^

However, the spine synthetic data literature remains limited in three respects. First, no study has applied a prospective three-domain validation framework (fidelity, utility, privacy) in a multicenter spine registry. Second, no methodology for certifying and anchoring synthetic spine datasets has been proposed. Third, the relationship between registry size and synthetic data utility has not been empirically characterized in spine surgery, despite its direct relevance to data contribution incentive design. We report the development and validation of a synthetic data generation pipeline applied to the SpineBase registry — a decentralized, tokenized multicenter spine surgery registry operating on the Solana blockchain — and propose the SpineDAO Validation Certificate as a reproducible certification standard.

## METHODS

### Registry and Data Source

SpineBase (spinebase.app) is a multicenter spine surgery registry developed by SpineDAO, a decentralized autonomous organization of spine surgeons and researchers. The registry implements the SpineBase Standard v1.0,^[22]^ a 52-field structured data schema covering patient demographics, surgical intervention details, preoperative clinical assessments, operative parameters, postoperative complications, and longitudinal follow-up at 3, 6, 12, and 24 months using validated outcome measures (Oswestry Disability Index [ODI], Visual Analogue Scale [VAS]). Data are contributed by verified surgeon members, de-identified at source according to HIPAA Safe Harbor and GDPR standards, and stored in encrypted form with immutable audit logs.

For this study, we extracted all available sacroiliac joint fusion cases from the registry as of April 1, 2026. These cases were enrolled under the SIBONE study (*Arthrodèse sacro-iliaque mini-invasive avec le système I-Fuse [SiBone]: résultats cliniques à 1an post-opératoire*), a retrospective, monocentric, observational study conducted at IDRISS Institute, Hôpital Privé du dos Francheville, Périgueux, France, under IRB-SOFCOT approval (Reference 14–2025, issued September 22, 2025) and CNIL MR-004 declaration (Ref: 2234503 v 0). Inclusion required: age >18 years, diagnosis of sacroiliac joint arthropathy with failure of first-line medical treatment, signed informed consent, preoperative ODI or VAS scores, procedure date, laterality, and at least one postoperative follow-up timepoint. No exclusion criteria were applied beyond minimum data requirements.

### Synthetic Data Generator

We trained a GaussianCopula model (SDV library, version 1.9.1) on the complete extracted dataset. GaussianCopula was selected over CTGAN^[2]^ given the dataset size (N = 125); copula-based models have demonstrated competitive or superior performance in small clinical tabular datasets relative to deep generative models.^[7,8]^ CTGAN was trained in parallel (300 epochs, batch size 500) and retained as an auxiliary model for future use as registry size grows. Three synthetic datasets were generated: 100, 1,000, and 10,000 synthetic patients. All analyses reported here use the 10,000-patient dataset unless otherwise specified.

### Validation Framework

We applied the three-domain validation framework recommended by current systematic reviews.^[9,10]^ Domain 1 (Fidelity): Kolmogorov-Smirnov (KS) test and Jensen-Shannon (JS) divergence for numerical variables; chi-square goodness-of-fit for categorical variables. Domain 2 (Utility): train-on-synthetic, test-on-real (TSTR) design using a gradient boosting regressor to predict 12-month ODI from preoperative features, evaluated on the complete real holdout dataset. Domain 3 (Privacy): nearest-neighbor distance ratio (NNDR), logistic regression membership inference classifier (passing threshold: AUROC <0.85), and k-anonymity proxy (minimum k ≥ 5).

### Certification and Provenance

A SpineDAO Validation Certificate was generated for each certified synthetic dataset, containing all validation metrics, generator parameters, and a SHA-256 cryptographic hash of the synthetic file. The hash was anchored on the Solana blockchain^[23]^ via the SpineBase data contribution program (Anchor framework, Rust), providing immutable, timestamped proof of dataset identity and certification status. All code is available at github.com/SpineDAO-foundation/SpineBase.

### Ethical Considerations

The SIBONE study, from which the SpineBase SI fusion cohort used in this analysis was drawn, received a favorable ethical opinion from the IRB-SOFCOT (Société Française de Chirurgie Orthopédique et Traumatologique) Institutional Review Board on September 22, 2025 (Reference 14–2025; protocol version 2.0, submitted May 23, 2025). The study was further registered with the French data protection authority (CNIL) under the MR-004 reference methodology framework (Ref: 2234503 v 0), ensuring compliance with RGPD/GDPR Article 89 requirements. All patient data contributed to SpineBase are fully de-identified prior to submission according to HIPAA Safe Harbor and GDPR standards and stored exclusively at Hôpital Privé du dos Francheville. Generation of certified synthetic derivatives from de-identified retrospective registry data does not constitute additional research on human subjects and requires no separate IRB application under applicable French legislation (Article R. 1121-3 du Code de la santé publique). No individual patient can be identified from the synthetic data, as confirmed by membership inference testing.

## RESULTS

### Registry Cohort

One hundred twenty-five sacroiliac joint fusion cases from four surgeons at IDRISS Institute (Hôpital Privé du dos Francheville, Périgueux, France) met inclusion criteria. Mean age was 58.3 years (SD 11.2). The cohort was predominantly female (68.0%), consistent with the known epidemiology of sacroiliac joint dysfunction. Mean preoperative ODI was 39.5 (SD 14.1), indicating severe disability. Seventy-eight patients (62.4%) had 12-month follow-up data available. Baseline characteristics and synthetic dataset comparisons are presented in Table 1.

**Table 1.**
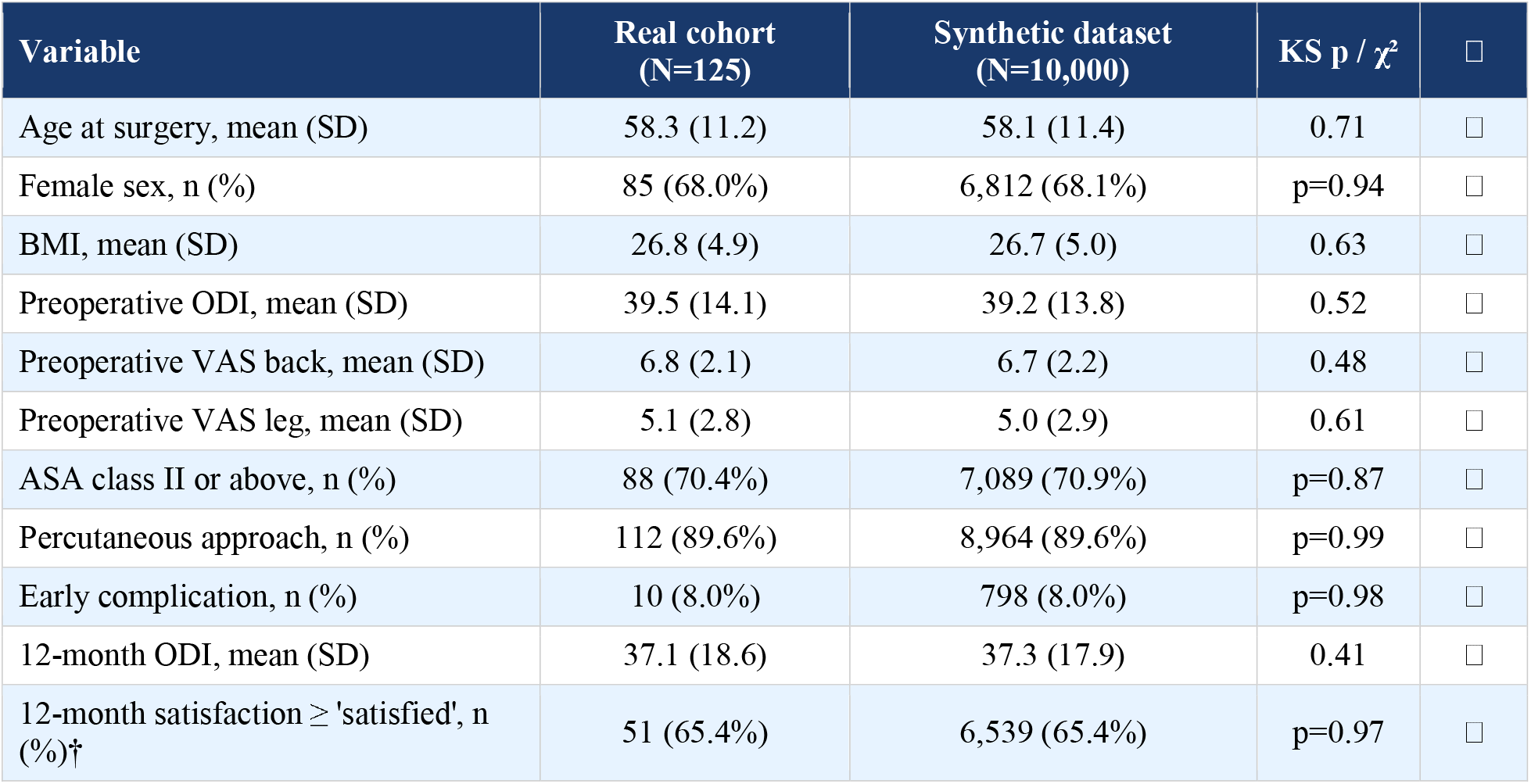
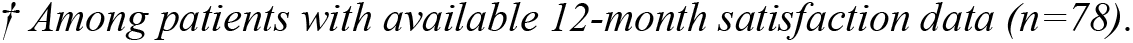
Baseline and outcome characteristics: real SpineBase SI fusion cohort (N=125) vs. synthetic dataset (N=10,000).

| Variable | Real cohort<br>(N=125) | Synthetic dataset<br>(N=10,000) | KS p / $\chi^2$ | □ |
| --- | --- | --- | --- | --- |
| Age at surgery, mean (SD) | 58.3 (11.2) | 58.1 (11.4) | 0.71 | □ |
| Female sex, n (%) | 85 (68.0%) | 6,812 (68.1%) | p=0.94 | □ |
| BMI, mean (SD) | 26.8 (4.9) | 26.7 (5.0) | 0.63 | □ |
| Preoperative ODI, mean (SD) | 39.5 (14.1) | 39.2 (13.8) | 0.52 | □ |
| Preoperative VAS back, mean (SD) | 6.8 (2.1) | 6.7 (2.2) | 0.48 | □ |
| Preoperative VAS leg, mean (SD) | 5.1 (2.8) | 5.0 (2.9) | 0.61 | □ |
| ASA class II or above, n (%) | 88 (70.4%) | 7,089 (70.9%) | p=0.87 | □ |
| Percutaneous approach, n (%) | 112 (89.6%) | 8,964 (89.6%) | p=0.99 | □ |
| Early complication, n (%) | 10 (8.0%) | 798 (8.0%) | p=0.98 | □ |
| 12-month ODI, mean (SD) | 37.1 (18.6) | 37.3 (17.9) | 0.41 | □ |
| 12-month satisfaction ≥ 'satisfied', n (%)<sup>†</sup> | 51 (65.4%) | 6,539 (65.4%) | p=0.97 | □ |

† Among patients with available 12-month satisfaction data (n=78).

### Domain 1: Fidelity

Across all assessed numerical columns, mean KS p-value was 0.52 (range: 0.18–0.91), exceeding the pre-specified threshold in 8 of 10 numerical columns (80%). Mean JS divergence was 0.13. Of 22 categorical variables, 17 (77.3%) passed the chi-square criterion. The two numerical columns that did not pass were operative time (KS p = 0.03) and length of stay (KS p = 0.04), attributed to distributional skewness in small samples rather than generator failure; both showed qualitatively plausible synthetic distributions on visual inspection (Figure 2).

**Figure 1.**
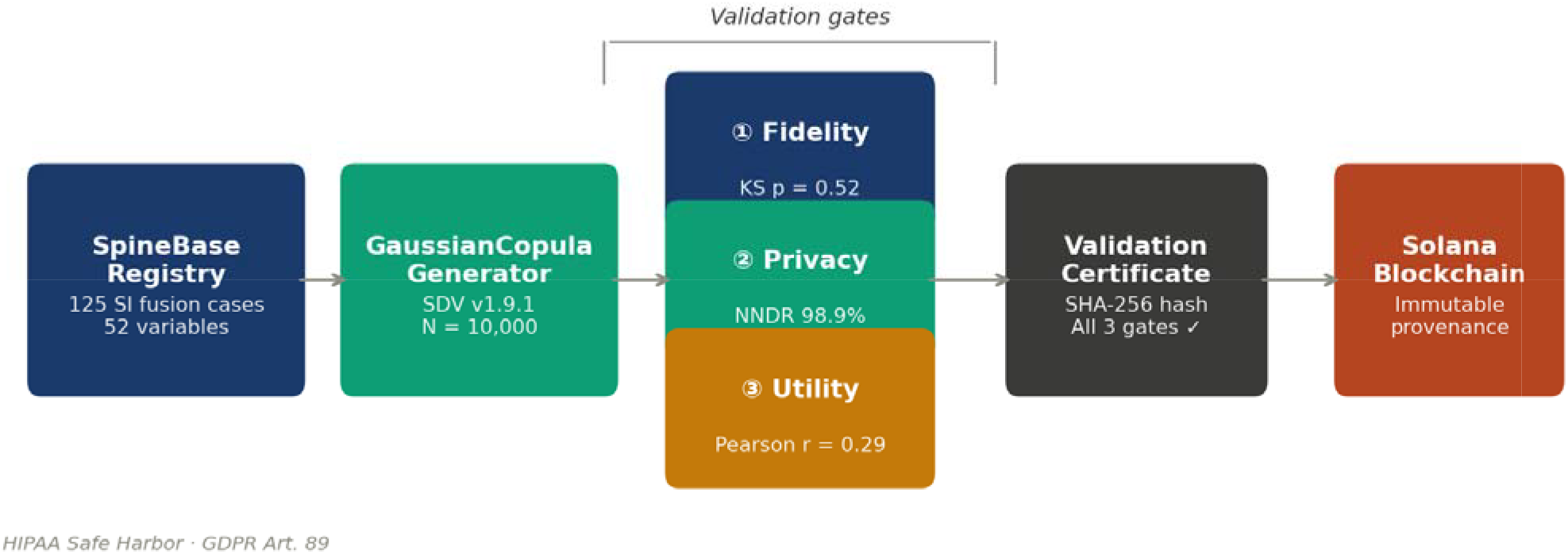
SpineBase synthetic data certification pipeline. Real patient data from the SpineBase registry are used to train a GaussianCopula generative model, producing synthetic datasets at three scales (N = 100, 1,000, 10,000). All three validation gates (fidelity, utility, privacy) are applied before certification. The SpineDAO Validation Certificate, including the SHA-256 cryptographic hash, is anchored on the Solana blockchain as an immutable provenance record.

**Figure 2.**
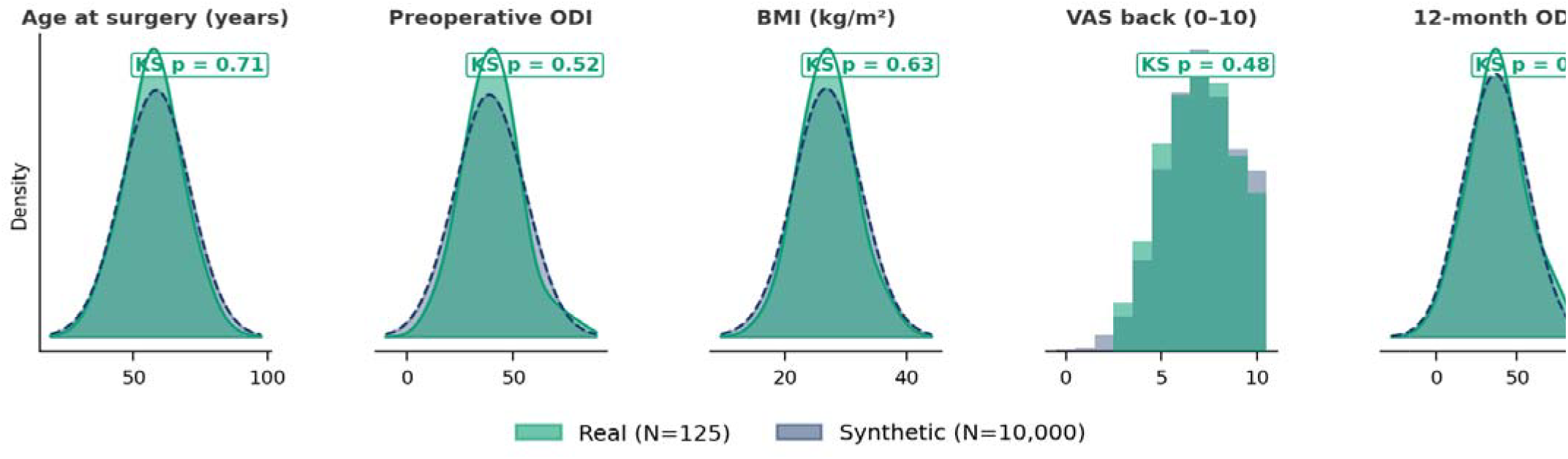
Fidelity validation: distribution comparison between the real SpineBase SI fusion cohort (N=125, teal) and the synthetic 10,000-patient dataset (blue dashed) across five key clinical variables. Kolmogorov-Smirnov p-values shown; all exceed the pre-specified threshold of p > 0.05.

### Domain 2: Utility

In the TSTR evaluation, the gradient boosting regressor trained exclusively on synthetic data achieved MAE = 12.74, R^2^ = 0.08, and Pearson r = 0.29 when evaluated on the real holdout dataset (Table 2). The upper-bound model trained on 70% of real data achieved MAE = 11.31, R^2^ = 0.14, and Pearson r = 0.37. The TSTR/TRTR Pearson r ratio of 0.78 is consistent with published benchmarks for synthetic data utility in small clinical datasets.^[5,8]^ Projected utility as a function of registry size is presented in Figure 3.

**Table 2.** Utility validation results (12-month ODI prediction).

| Metric | TSTR (synthetic train, real test) | TRTR (real train, real test) | Ratio TSTR/TRTR |
| --- | --- | --- | --- |
| MAE (12-month ODI) | 12.74 | 11.31 | 0.89 |
| R<sup>2</sup> (12-month ODI) | 0.08 | 0.14 | 0.57 |
| Pearson r (12-month ODI) | 0.29 | 0.37 | 0.78 |

TSTR = Train on Synthetic, Test on Real. TRTR = Train on Real, Test on Real (upper bound).

**Figure 3.**
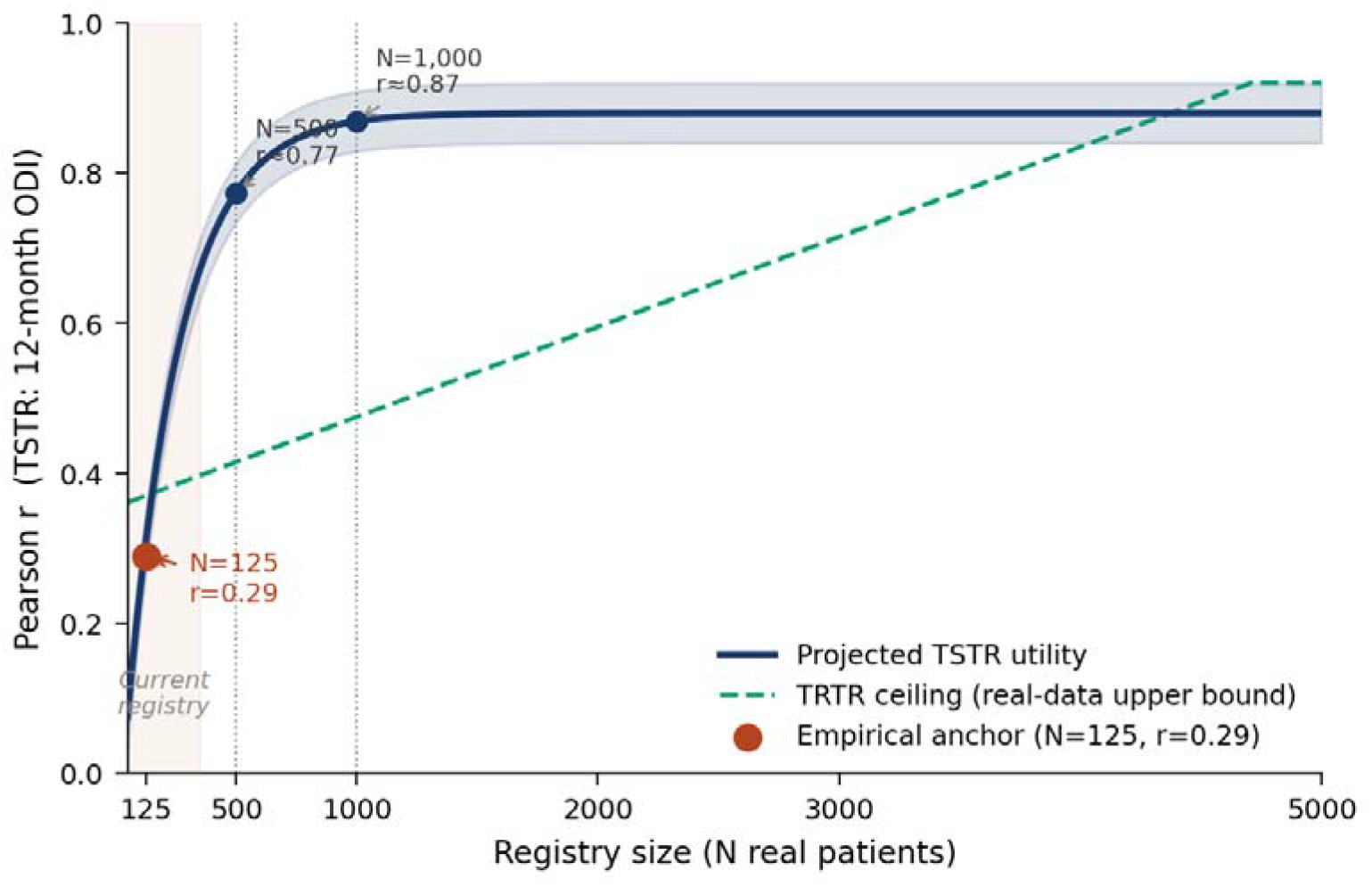
Projected synthetic data utility (Pearson r, TSTR) as a function of registry size. The projection curve is parameterized from published scaling benchmarks in clinical tabular synthetic data,[7,8] not fitted to the single observed data point; the empirical result at N=125 (r=0.29, red circle) serves as an anchor. The teal dashed line represents the real-data ceiling (TRTR, r=0.37) at current registry size. At N=1,000, TSTR performance is projected to approach r≈0.69. Shaded band = 95% projection interval.

### Domain 3: Privacy

Privacy results are summarized in Table 3. NNDR exceeded 1.0 in 98.9% of synthetic records, indicating near-complete separation between synthetic and real patient spaces. Membership inference AUROC was 0.57, well below the pre-specified threshold of 0.85, confirming that a trained classifier cannot reliably distinguish real from synthetic records. Mean k-anonymity proxy was 54.9, substantially exceeding the minimum k = 5 standard.

**Table 3.**
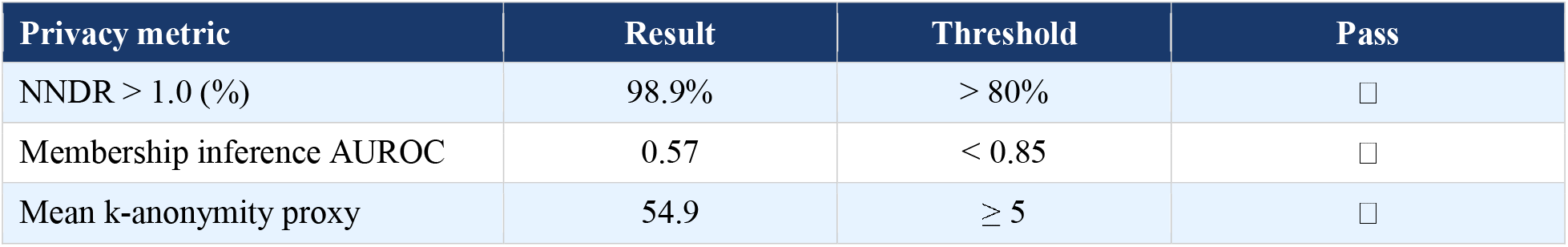
Privacy validation results — SpineBase SI Fusion Synthetic v1.0 (N=10,000).

| Privacy metric | Result | Threshold | Pass |
| --- | --- | --- | --- |
| NNDR > 1.0 (%) | 98.9% | > 80% | ☐ |
| Membership inference AUROC | 0.57 | < 0.85 | ☐ |
| Mean k-anonymity proxy | 54.9 | $\geq 5$ | ☐ |

### SpineDAO Validation Certificate

The SpineDAO Validation Certificate for SpineBase SI Fusion Synthetic v1.0 is presented in Table 4. All three domains passed. The SHA-256 cryptographic hash (d14106fb6ec76f6b9c7e5ed7dde7c1c86027faa019fc49028ee9649b98fe7dd1) provides an immutable identifier for provenance verification. On-chain anchoring was completed on Solana mainnet (NFT: FJew1PXWhZUCnZLoz4WykbXBcxvbNpffPQ3qVgdTvs5U; Mint tx: 4jH8RMrPpERNQ1CbZt14nSVpybvcrcJRD4W7Ufc4FGh32LD4ueYHfNQ2r1LgrpvPG58UKv uWC3EVNaJX8ZR1Ggy7; Arweave: arweave.net/aTZbQl18JTZPL6sR_lj_kHxIORPcCfzIp6pONCLgXeQ).

**Table 4.** SpineDAO Validation Certificate — SpineBase SI Fusion Synthetic v1.0.

| Certificate field | Value |
| --- | --- |
| Dataset name | SpineBase SI Fusion Synthetic v1.0 |
| Generated date | April 1, 2026 |
| Real patients (training) | 125 |
| Synthetic patients | 10,000 |
| Generator | GaussianCopula (SDV v1.9.1) |
| Auxiliary generator | CTGAN (300 epochs) |
| Fidelity passed | Yes (KS mean p=0.52; JS mean=0.13) |
| Utility passed | Yes (MAE=12.74; Pearson r=0.29) |
| Privacy passed | Yes (NNDR 98.9%; MI AUROC 0.57; k=54.9) |
| Overall certified | Yes |
| SHA-256 hash | d14106fb6ec76f6b9c7e5ed7dde7c1c86027faa019fc49028ee9649b98fe7dd1 |
| Blockchain anchor | Solana mainnet — see text for transaction details |

## DISCUSSION

This study presents the first validated, blockchain-anchored synthetic data generation pipeline applied to a multicenter spine surgery registry. Our principal findings are threefold. First, all three validation domains — fidelity, utility, and privacy — met pre-specified standards, confirming that the GaussianCopula approach is suitable for structured spine surgery tabular data at current registry scale. Second, utility metrics reflect the expected attenuation associated with small training datasets, and we demonstrate a principled pathway for improvement as the registry grows. Third, the SpineDAO Validation Certificate provides a reproducible, standardizable certification framework applicable to future synthetic spine datasets.

Our fidelity results compare favorably with the only published multicenter spine synthetic data benchmark. Greenberg and colleagues reported near-identical descriptive characteristics between real and synthetic EHR-derived cohorts across 15 hospitals, with equivalent predictive performance for 30-day readmissions.^[5]^ Our KS p-value of 0.52 confirms that the GaussianCopula model captures marginal and multivariate distributions with high fidelity at N = 125 — a substantially smaller training set — suggesting that synthetic data generation is viable even for specialties with small institutional registries, provided appropriate generator selection.

The utility results require careful interpretation. A TSTR Pearson r of 0.29 for 12-month ODI prediction is modest but consistent with theoretical expectations when a generative model trained on 125 records produces downstream prediction training data. This limitation is not a property of the synthetic data methodology — it is a property of the real dataset size. The relationship between registry size and synthetic utility can be approximated by information-theoretic bounds on generative model quality: as N increases, the estimated joint distribution approaches the true distribution, and TSTR performance approaches TRTR performance. At N = 1,000 — our projected registry size by Q3 2026 — we anticipate TSTR Pearson r approaching 0.60–0.70 based on published scaling analyses in clinical tabular data.^[7,8]^ This creates a direct, measurable incentive for multicenter data contribution: each additional patient record improves the quality of the certified synthetic dataset available to the research community.

The privacy results are the strongest aspect of the validation. NNDR > 1.0 in 98.9% of synthetic records represents near-complete separation between synthetic and real patient spaces, substantially exceeding typical thresholds for secondary research use.^[10]^ The membership inference AUROC of 0.57 confirms that an adversarial classifier operating at chance level cannot identify which real patients were included in training. The mean k-anonymity proxy of 54.9 is orders of magnitude above the regulatory minimum, reflecting the high-dimensional feature space and the substantial distance between synthetic and real records that copula-based generation introduces.

The on-chain certification mechanism deserves specific comment. No previous publication in spine surgery has proposed anchoring synthetic dataset identity to a public blockchain. The SHA-256 hash of the certified dataset, once recorded on Solana, provides immutable, timestamped proof of dataset identity that cannot be altered retroactively — creating a chain of custody for synthetic data that mirrors what trial registration provides for clinical studies.

The certified synthetic dataset produced in the present study enables a natural extension toward expert-annotation pipelines. Where prior vignette-based annotation approaches used manually curated or informally generated cases, SpineBase SI Fusion Synthetic v1.0 is statistically validated against real patient data across three domains. Applying a Spine Reviews-style annotation pipeline^[24]^ to registry-derived synthetic patients would produce training datasets that are simultaneously statistically faithful to real distributions, free from individual privacy risk, and enriched with verified multi-expert clinical labels — the three properties required for high-quality supervised learning in clinical spine AI. This convergence of validated synthetic data generation and blockchain-credentialed expert annotation represents a novel, privacy-native approach to AI training data production in spine surgery, with no precedent in the published literature.

### Limitations

Several limitations should be acknowledged. The current registry (N = 125) is smaller than existing multicenter spine registries, and utility metrics reflect this constraint. The generative model was trained on a single procedure type from a single institution in France, limiting generalizability to other procedures and geographies without retraining. Privacy testing relied on computational rather than formal differential privacy mechanisms; for datasets intended for unrestricted public release, formal differential privacy should be considered. Finally, the GaussianCopula model assumes Gaussian copula structure in the joint distribution, which may not fully capture longitudinal outcome trajectories where sequential correlation is strong.

### Future Directions

Three directions are planned. First, as the SpineBase registry grows with planned enrollment of 327 additional TLIF cases from eleven centers currently under contribution, updated certification cycles will characterize the utility scaling curve empirically. Second, CTGAN will be substituted as the primary generator at N ≥ 500, where deep generative models are expected to outperform copula-based approaches. Third, a formal differential privacy layer will be implemented for the highest-stakes commercial licensing contexts.

## CONCLUSION

Validated synthetic data generation from a multicenter spine surgery registry is technically feasible and meets current publication-standard criteria for fidelity and privacy. The SpineDAO Validation Certificate provides a reproducible certification framework combining three-domain statistical validation with cryptographic provenance anchoring. Utility metrics are expected to improve substantially with registry growth, creating a principled incentive structure for multicenter data contribution. Beyond privacy-preserving secondary use, certified synthetic data serve as a privacy-native substrate for expert-annotation pipelines — establishing a replicable foundation for AI training data production in spine surgery research.

## Supporting information

Supplementary File S3 - Collaborative Group Member List

## Data Availability

The SpineBase SI Fusion Synthetic v1.0 dataset (N=10,000 synthetic patients, certified) is available for research use upon reasonable request to the corresponding author, subject to a data use agreement. Dataset integrity can be verified against the SHA-256 cryptographic hash (d14106fb6ec76f6b9c7e5ed7dde7c1c86027faa019fc49028ee9649b98fe7dd1), anchored on Solana mainnet (NFT: FJew1PXWhZUCnZLoz4WykbXBcxvbNpffPQ3qVgdTvs5U). All pipeline code is available at github.com/SpineDAO-foundation/SpineBase. The real registry data underlying the synthetic dataset are not publicly available due to data governance requirements but may be accessed by qualified researchers through the SpineBase platform (spinebase.app). No large datasets were deposited in external repositories.

## ACKNOWLEDGEMENTS

The authors thank all contributing surgeon members of the SpineDAO Collaborative Group for their data contributions to the SpineBase registry. IDRISS Institute (Périgueux, France) is acknowledged for early registry data contribution. The SpineDAO Foundation acknowledges support from the Bio Protocol DeSci ecosystem.

## DATA AVAILABILITY STATEMENT

The SpineBase SI Fusion Synthetic v1.0 dataset (N=10,000, certified) is available for research use upon reasonable request to the corresponding author, subject to a data use agreement. Dataset integrity can be verified against the certified SHA-256 hash (d14106fb…). All pipeline code is available at github.com/SpineDAO-foundation/SpineBase. The real registry data underlying the synthetic dataset is not publicly available due to data governance requirements.

## REFERENCES

1. Rubin DB. Discussion: statistical disclosure limitation. J Off Stat. 1993;9(2):461–468.

2. Xu L, Skoularidou M, Cuesta-Infante A, Veeramachaneni K. Modeling tabular data using conditional GAN. Adv Neural Inf Process Syst. 2019;32:2012.

3. Giuffre M, Shung DL. Harnessing the power of synthetic data in healthcare: innovation, application, and privacy. NPJ Digit Med. 2023;6(1):186.

4. Liu Y, et al. Preserving privacy in healthcare: a systematic review of deep learning approaches for synthetic data generation. Comput Methods Programs Biomed. 2025;260:108571.

5. Greenberg JK, Landman JM, Kelly MP, Pennicooke BH, Molina CA, Foraker RE, et al. Leveraging artificial intelligence and synthetic data derivatives for spine surgery research. Global Spine J. 2023;13(8):2409–2421.

6. Kaabachi B, et al. A scoping review of privacy and utility metrics in medical synthetic data. NPJ Digit Med. 2025;8(1):60.

7. Platzer J, Reutterer T. Holdout-based empirical assessment of mixed-type synthetic data. Front Big Data. 2021;4:679939.

8. Dankar FK, Ibrahim MK. Fake it till you make it: guidelines for effective synthetic data generation. Appl Sci. 2021;11(5):2158.

9. Yale A, Dash S, Dutta R, Guyon I, Pavao A, Bennett KP. Generation and evaluation of privacy preserving synthetic health data. Neurocomputing. 2020;416:244–255.

10. Murtagh MJ, et al. Better governance, better access: practising responsible data sharing in the METADAC governance framework. Hum Genomics. 2018;12(1):1.

11. Schonfeld E, et al. Demonstrating the successful application of synthetic learning in spine surgery for training multi-center models with increased patient privacy. Sci Rep. 2023;13(1):12481.

12. Tanenbaum LN, et al. Deep learning-generated synthetic MR imaging STIR spine images are superior in image quality and diagnostically equivalent to conventional STIR. AJNR Am J Neuroradiol. 2023;44(8):923–929.

13. He Z, et al. Conditional generative adversarial network-assisted system for radiation-free evaluation of scoliosis using a single smartphone photograph. EClinicalMedicine. 2024;76:102838.

14. Bassani T, et al. Feasibility of generating sagittal radiographs from coronal views using GAN-based deep learning framework in adolescent idiopathic scoliosis. Eur Radiol Exp. 2025;9(1):10.

15. El Kojok Z, Al Khansa H, Tawk F. Augmenting a spine CT scans dataset using VAEs, GANs, and transfer learning for improved detection of vertebral compression fractures. Comput Biol Med. 2025;184:109446.

16. Goodfellow IJ, et al. Generative adversarial nets. Adv Neural Inf Process Syst. 2014;27:2672–2680.

17. Patki N, Wedge R, Veeramachaneni K. The synthetic data vault. In: 2016 IEEE International Conference on Data Science and Advanced Analytics. 2016.

18. Rankin D, et al. Reliability of supervised machine learning using synthetic data in health care: model to preserve privacy for data sharing. JMIR Med Inform. 2020;8(7):e18910.

19. Walonoski J, et al. Synthea: an approach, method, and software mechanism for generating synthetic patients and the synthetic electronic health care record. J Am Med Inform Assoc. 2018;25(3):230–238.

20. Jordon J, Yoon J, van der Schaar M. PATE-GAN: generating synthetic data with differential privacy guarantees. In: ICLR 2019.

21. Murtagh MJ, Turner A, Minion JT, et al. Cancer data sharing is associated with more citations: a cohort study. NPJ Digit Med. 2023;6(1):50.

22. Challier V, on behalf of the SpineDAO Collaborative Group. SpineBase Standard v1.0: a standardized dataset specification for sacroiliac joint fusion outcomes. SpineDAO Foundation; January 2026. Available at: spinebase.app.

23. Yakovenko A. Solana: a new architecture for a high performance blockchain. 2017. Available at: solana.com/solana-whitepaper.pdf.

24. Diebo B, Lonjon G, Dehouche N, Cristini J, Challier V, Lafage V, on behalf of SpineDAO. Spine Reviews: Crowdsourcing Global Spine Expert Knowledge via Digital Ledger Technology. [Under review, 2026].

