## Supplementary File S3 - Collaborative Group Member List for "Validated Synthetic Data Generation from a Multicenter Spine Surgery Registry: Methodology and Benchmark"

**Supplementary Appendix S3**

**SpineDAO Collaborative Group — Complete Member List**

*Validated Synthetic Data Generation from a Multicenter Spine Surgery Registry: Methodology and Benchmark*

*Challier V, Jacquemin C, Diebo B, Dehouche N, Denisov A, Cristini J, Campana M, Castelain J-E, Lonjon G, Lafage V, Ghailane S, on behalf of the SpineDAO Collaborative Group. medRxiv, 2026.*

The SpineDAO Collaborative Group comprises verified spine surgeons, neurosurgeons, and clinical researchers contributing outcomes data, governance, or scientific expertise to the SpineBase multicenter registry. Membership is credentialed via Soulbound Tokens (SBTs) on the Solana blockchain — non-transferable digital credentials permanently bound to each member's verified professional identity. Members listed as 'Pending' hold confirmed credentials; SBT minting is deferred for technical reasons unrelated to credential status. All members have consented to inclusion in this appendix.

| **Name** | **Degree** | **Role / Specialty** | **Institution** | **Country** | **SBT** |
| --- | --- | --- | --- | --- | --- |
| **Vincent Challier** | MD | Spine Surgeon · Founder | Hôpital Privé du dos Francheville (IDRISS Institute) | France | ✓ Minted |
| **Clément Jacquemin** | PhD | Study Coordinator | Hôpital Privé du dos Francheville (IDRISS Institute) | France | — |
| **Matthieu Campana** | MD | Spine Surgeon | Hôpital Privé du dos Francheville (IDRISS Institute) | France | ✓ Minted |
| **Jean-Etienne Castelain** | MD | Spine Surgeon | Hôpital Privé du dos Francheville (IDRISS Institute) | France | — |
| **Soufiane Ghailane** | MD | Spine Surgeon · SIBONE PI | Hôpital Privé du dos Francheville (IDRISS Institute) | France | — |
| **Mathieu Vassal** | MD | Spine Surgeon | France | France | ✓ Minted |
| **David Giber** | MD | Spine Surgeon | France | France | ✓ Minted |
| **Alexis Perez** | MD | Spine Surgeon | France | France | ✓ Minted |
| **B. Liabaud** | MD | Spine Surgeon | France | France | ✓ Minted |
| **Jérôme Delambre** | MD | Spine Surgeon | France | France | ✓ Minted |
| **Alexandre Delmotte** | MD | Spine Surgeon | France | France | ✓ Minted |
| **Alexandre Dhenin** | MD | Spine Surgeon | France | France | Pending |
| **Guillaume Lonjon** | MD | Spine Surgeon | Clinique Orthosud, Saint Jean de Védas | France | Pending |
| **Joseph Cristini** | MD | Neurosurgeon | Hôpital Clairval, Marseille | France | — |
| **Bassel Diebo** | MD | Orthopaedic Spine Surgeon | Brown University, Dept. of Orthopaedic Surgery, Providence RI | USA | — |
| **Virginie Lafage** | PhD | Spine Biomechanics Researcher | Lenox Hill Hospital, Dept. of Orthopaedic Surgery, New York NY | USA | Pending |
| **Anton Denisov** | MD, PhD | Spine Surgeon · Biostatistician | University Hospital Mollet, Dept. of Orthopedic Surgery, Barcelona / The Taylor Collaboration, San Francisco CA | Spain / USA | ✓ Minted |
| **Nassim Dehouche** | PhD | Researcher | Mahidol University International College, Salaya | Thailand | Pending |

**SBT status as of April 7, 2026.** ✓ Minted = Soulbound Token confirmed on Solana mainnet. Pending = credentials verified; minting deferred for technical reasons. — = non-SBT contributor (data contributor, study coordinator, or co-investigator). The 'SpineDAO Approved Clinicians' SBT collection is deployed on Solana mainnet. SBTs encode specialization, country of practice, and token type on-chain and are non-transferable by design.

SpineDAO Collaborative Group governance is managed via the SpineDAO Foundation DAO. Membership decisions, data contribution standards, and research grant allocations are subject to token-weighted governance votes by $SPINE holders. Further information: spinal.science · spinebase.app · spinedao.com
